# Finding a Needle in a Haystack: Design and Implementation of a Digital Site-less Clinical Study of Serial Rapid Antigen Testing to Identify Asymptomatic SARS-CoV-2 Infection

**DOI:** 10.1101/2022.08.04.22278274

**Authors:** Apurv Soni, Carly Herbert, Caitlin Pretz, Pamela Stamegna, Andreas Filippaios, Qiming Shi, Thejas Suvarna, Emma Harman, Summer Schrader, Chris Nowak, Eric Schramm, Vik Kheterpal, Stephanie Behar, Seanan Tarrant, Julia Ferranto, Nathaniel Hafer, Matthew Robinson, Chad Achenbach, Robert L. Murphy, Yukari C. Manabe, Laura Gibson, Bruce Barton, Laurel O’Connor, Nisha Fahey, Elizabeth Orvek, Peter Lazar, Didem Ayturk, Steven Wong, Adrian Zai, Lisa Cashman, Lokinendi V Rao, Katherine Luzuriaga, Stephenie Lemon, Allison Blodgett, Elizabeth Trippe, Mary Barcus, Brittany Goldberg, Kristian Roth, Timothy Stenzel, William Heetderks, John Broach, David McManus

## Abstract

**Background:** Rapid antigen tests (Ag-RDT) for SARS-CoV-2 with Emergency Use Authorization generally include a condition of authorization to evaluate the test’s performance in asymptomatic individuals when used serially.

**Objective:** To describe a novel study design to generate regulatory-quality data to evaluate serial use of Ag-RDT in detecting SARS-CoV-2 virus among asymptomatic individuals.

**Design:** Prospective cohort study using a decentralized approach. Participants were asked to test using Ag-RDT and molecular comparators every 48 hours for 15 days.

**Setting:** Participants throughout the mainland United States were enrolled through a digital platform between October 18, 2021 and February 15, 2022. Ag-RDTs were completed at home, and molecular comparators were shipped to a central laboratory.

**Participants:** Individuals over 2 years old from across the U.S. with no reported COVID-19 symptoms in the 14 days prior to study enrollment were eligible to enroll in this study.

**Measurements:** Enrollment demographics, geographic distribution, and SARS-CoV-2 infection rates are reported.

**Key Results:** A total of 7,361 participants enrolled in the study, and 492 participants tested positive for SARS-CoV-2, including 154 who were asymptomatic and tested negative to start the study. This exceeded the initial enrollment goals of 60 positive participants. We enrolled participants from 44 U.S. states, and geographic distribution of participants shifted in accordance with the changing COVID-19 prevalence nationwide.

**Limitations:** New, complex workflows required significant operational and data team support. Conclusions: The digital site-less approach employed in the ‘Test Us At Home’ study enabled rapid, efficient, and rigorous evaluation of rapid diagnostics for COVID-19, and can be adapted across research disciplines to optimize study enrollment and accessibility.

## INTRODUCTION

Over-the-counter rapid antigen SARS-CoV-2 tests (Ag-RDT) present unique opportunities for widespread COVID-19 testing. However, the authorization of these tests under the Food and Drug Administration’s (FDA) Emergency Use Authorization (EUA) generally imposes a requirement on companies to evaluate their test’s performance for detecting asymptomatic infections with serial screening. To satisfy this requirement, it was necessary to conduct a longitudinal study to observe the onset of SARS-CoV-2 infection and performance of Ag-RDT across the course of infection.^5^ This required an innovative research design to generate regulatory-quality, representative data, capture new-onset infections amidst a changing pandemic, and understand asymptomatic infections, which represent a small proportion of all SARS-CoV-2 cases.

This manuscript describes the design, implementation, and completion of a novel, digital, site-less study called ‘Test Us at Home’ funded by the National Institutes of Health (NIH) Rapid Acceleration Diagnostics (RADx) Program. In this manuscript, we outline the rationale for key decision-making and describe findings of the operational effort needed to successfully collect data under this paradigm, to provide rapid, rigorous evaluation of over-the-counter diagnostics across diverse participants of all ages in community-based settings.

## METHODS

### Study Design

This prospective cohort study recruited participants over two-years old who were asymptomatic and self-reported absence of SARS-CoV-2 infection in the three months prior to enrollment. Participants were asked to test for SARS-CoV-2 via Ag-RDT and molecular comparator every 48-hours for 15 days. This study was approved by the Institutional Review Board of the WIRB-Copernicus Group.

### Study Objectives

The primary objective of this collaboration between the FDA, NIH, and the RADx Tech Clinical Studies Core at UMass Chan Medical School was to generate right-of-reference data that could be used towards FDA EUA. To accomplish this, we aimed to evaluate the performance of serial use of three different Ag-RDT to detect a new-onset SARS-CoV-2 infection, as determined by a laboratory-based molecular test.

#### Study Population and Recruitment

To approximate the performance of Ag-RDT in the general public, the FDA suggested against preferentially enrolling participants with a known SARS-CoV-2 exposure (oral communication, August 3, 2021). The goal of the study was to recruit at least 60 participants who started the study without any symptoms and with a negative SARS-CoV-2 molecular test and subsequently tested positive during the study period. Due to the fluctuating rates of COVID-19 positivity and the importance of recruiting asymptomatic participants, we employed a decentralized study design whereby eligible participants from anywhere in the mainland United States except for Arizona could participate. Residents from the state of Arizona were excluded because Quest Diagnostics, the laboratory performing molecular tests, did not provide direct-to-consumer tests in that state. Additional inclusion and exclusion criteria are listed in Supplemental Table 1. Recruitment relied on the engagement of community stakeholders to advertise the study and reach communities nationwide. Periodic analyses were conducted to inform enrollment goals based on observed precision for positive percent agreement.

#### Enrollment

Participants who expressed interest in the study received an email or flyer with instructions for downloading the study app and performing eligibility screening. The study app was a custom interface designed through the MyDataHelps platform. To enroll, participants answered a series of questions to determine their eligibility and prioritization status. All eligible participants underwent automated prioritization based on a-priori criteria that were adjusted on a daily or weekly basis (Supplemental Figure 1, Supplemental Figure 2). We adjusted prioritization criteria to preferentially enroll populations based on their region’s SARS-CoV-2 transmission rates, community-level vaccination, and sociodemographic characteristics. This step occurred prior to consenting, and data was discarded if the participant did not enroll in the study before the data collection period ended; these data were never accessible to the study team unless the participant consented to take part in the study. Eligible participants who did not match the preferred criteria for enrollment received a note saying that they cannot enroll in the study at this time but would receive communication from the study team when/if they became eligible (Supplemental Figure 3). These participants were placed on a “waiting list”, which was reviewed with regard to changing priority criteria throughout the study.

If eligible for enrollment, participants 18 years of age or older were asked to read the e-consent form and provide their signature through the study app (Supplemental Figure 4). Participants younger than 18 years were asked to review the e-assent and e-consent form with the support of their parent(s) or guardian(s) and provide assent for participation, while the parent/guardian provided written consent for participation in the study. Participants were eligible to receive up to $250 for timely completion of all sample collections (Supplemental Figure 5).

#### Test Distribution

On enrollment, participants were asked to provide their shipping information through the study app. Participants were assigned to one of three Ag-RDT (Quidel QuickVue At-Home OTC COVID-19 Test, Abbott BinaxNOW COVID-19 Antigen Self Test, or BD Veritor™ At-Home COVID-19 Test) using an automated algorithm that was informed by the investigators’ discretion based on enrollment numbers and geographic location of the participants (Supplemental Figure 6). The three Ag-RDT used in the study are authorized for emergency use by the FDA and were selected to provide multiple assessments and facilitate generalizing results to non-participating companies. These tests will be hereby referred to as test brands A, B, and C (randomly assigned), as the purpose of the study was not to directly compare different test performances. Participants residing together and enrolling with the same address or enrolled in a group, such as a cohort of classmates, were assigned the same Ag-RDT, to avoid mixing of test types between participants. Groups were given unique codes to input on enrollment, to ensure members were easily identifiable. As a result of this strategy, the assignment of tests to participants was non-random, and we intentionally did not pursue a strategy to compare performance between different tests.

Because molecular assays require a prescription by a licensed physician, the study team contracted with PWNHealth, a national clinician network, and developed an automated order filing system with Quest Diagnostics to place the requisite orders (Supplemental Figure 7). A total of 10 Ag-RDT and 7 home-collection kits for molecular comparators were provided to each participant. Participants received Ag-RDT and PCR home-collection kits by mail through separate shipments. The study team provided additional tests to participants if needed (e.g. tests were damaged upon delivery or lost in the mail).

#### Testing Schedule

Participants were asked to perform the Ag-RDT and collect the specimen for molecular comparator testing on the same day every 48 hours during the 15-day testing period (Supplemental Figure 8). Immediately prior to testing, participants were asked to record any symptoms on the day of testing. The study app provided a push-notification testing reminder at the 44-hour mark and additional notifications every two hours until the 52-hour mark if the participant had not completed their tests. Participants were instructed via the app to have at least a 15-minute break between the Ag-RDT and the sample collection for molecular tests. Participants were asked to record the results of the Ag-RDT when available by selecting one of the following options: “Negative”, “Positive”, “Invalid”, or “Don’t Know”, and upload a photo of the test strip through the app (Supplemental Figure 9). Participants assigned to Test C were also asked to download the Test C company-specific app which contained a test reader. The company test reader asked participants to upload a photo of the test strip and provided Ag-RDT results in real time. Test C users were asked to report the test result given by the test reader, rather than self-interpretation, to the study app. If the participant tested positive by Ag-RDT, they were told to contact their healthcare provider for any medical questions. In the event of an invalid Ag-RDT result, the participant was asked to perform a second Ag-RDT. If the second Ag-RDT result was also invalid, the participant was not asked to perform additional Ag-RDT on that day and was instructed to continue to the PCR sample collection (at least 15 minutes after Ag-RDT sample collection). Participants were responsible for shipping the PCR collection kit containing the sample using the pre-paid FedEx envelope using instructions provided by Quest Diagnostics, which were authorized by the FDA for emergency use.

#### Molecular Testing Procedures

Due to concerns of potential false-positive molecular tests, the FDA recommended the use of two types of molecular assays, and potentially, a third assay, if the prior two assays were discordant (oral communication, August 3, 2021).^2,3,4^ Once participant samples were received at Quest Diagnostics, the sample was divided into aliquots to perform Roche Cobas 6800 SARS-CoV-2 PCR and Quest RC COVID-19 PCR DTC for all participants (Supplemental Figure 10). An additional aliquot was preserved by the laboratory to allow for testing on Hologic Aptima SARS-CoV-2 Transcription Mediated Amplification (TMA) assay if Roche Cobas and Quest LDT were discordant or inconclusive. All three molecular assays used were highly sensitive, authorized by EUA, and run per instructions per use. Remnant samples of sample fluid from home-collection kits were frozen at -80 Celsius and shipped to UMass Chan Medical School for future testing. Results of the molecular PCR assays were available to participants through the Quest results portal and communication from the study team. All participants that tested positive by molecular test received a phone call from a study team member. In the event of discordant results, the participant was contacted by the study staff to explain the results.

#### Communication with the study team

Participants were provided a study hotline staffed with research coordinators to provide support during extended hours during the weekdays from 8 AM to 9 PM EST. Participants were also able to contact the study team via email at all times. All calls between coordinators and participants were documented in a call log containing the length of the call, the reason for the call, narrative about the call, and whether the issue was resolved.

#### Data Management

There were three primary sources of data (Supplemental Figure 11). All participant-reported data were collected through the study app and downloaded incrementally through secure file transfer protocol to UMass Chan servers. The molecular testing data were shared by Quest Diagnostics through an established datafeed between Quest and UMass Chan servers. However, the PCR cycle threshold (CT) values and results of the tiebreaker assays were not part of routinely abstracted data and required manual abstraction. Finally, tracking reports were used to document all communications between participants and the study staff. All three sources of data were combined using a participant’s unique identifier assigned by the study app. Paired Ag-RDT and PCR data were merged by matching on participant identifier and date of testing. When test dates were not aligned or there were unmatched results, a member of the study team reviewed all testing data from the participants to adjudicate paired findings. The most common reason for mismatched dates was due to missing dates of collection or transcription errors, as date of collection for PCR sample was handwritten by the participant on the Quest requisition form. Study staff reconciled possible mismatched test results by reviewing data for requisition numbers entered in the app, shipment tracking data, and contact reports. If mismatched data could not be adequately aligned, those data were considered ineligible due to failed quality-checks. Additional adjudication undertaken by the study team included a manual review of all Ag-RDT images for participants who either had a self-reported positive, don’t know, or invalid Ag-RDT result or if they tested positive on molecular test at any point during the study. All data were de-identified and shared with the FDA throughout the study. At the end of the study, the final and cleaned dataset was shared with the FDA and company-specific data were shared with the companies. All data will be made available on the NIH Data Hub once the main findings from this study are published.

## RESULTS

### Enrollment

In total, 7,361 participants enrolled in the study between October 18, 2021 and February 15, 2022 (Figure 1a). Due to complications with the company app, Test C enrollment began on November 4, 2021, three weeks after Test A and B enrollment started (Figure 1b). Following initial surges in recruitment, the enrollment waitlist was implemented on November 24, 2021 to allow for refinement of enrollment from geographic hotspots. In total, 492 participants tested positive on either molecular or Ag-RDT assays during the study, and 154 were eligible for analyses (Supplemental Figure 12).

**Figure 1:**
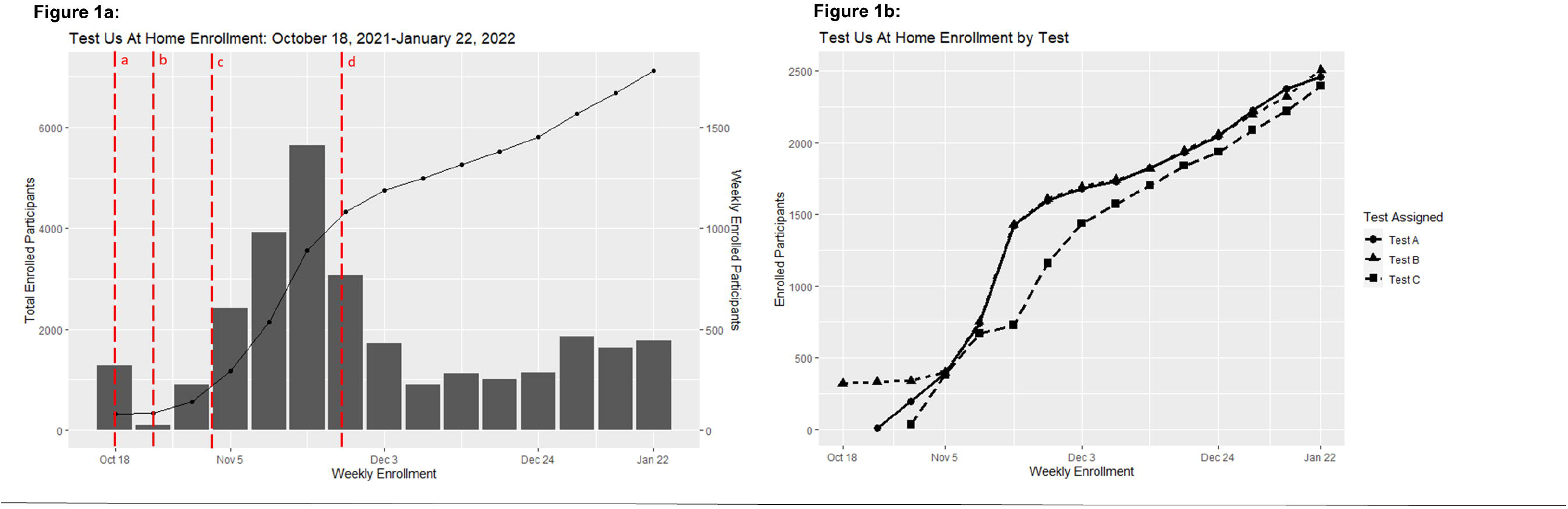
Test Us At Home Study Enrollment. a: Test A and B enrollment opens to individuals with specific group codes on October 18, 2021; b: Enrollment opens nationwide on October 26, 2021; c: Test C enrollment starts on November 4, 2021; d: Enrollment waitlist approach implemented on November 24, 2021

### Geographic and Sociodemographic Participant Characteristics

The geographic and sociodemographic characteristics of study participants changed over the course of the study, reflecting the shifts in the enrollment approach. In total, participants from 44 of the 48 mainland U.S. states enrolled in the study (Figure 2). In October, the majority of initial study recruitment occurred on a college campus in Wisconsin, resulting in a young, predominantly white, student population (Figure 2; Table 1). Additionally, only 3.1% of enrolled participants in October were unvaccinated for SARS-CoV-2. In November, study recruitment was expanded throughout the United States, with enrollment from more than 20 different states. Participants from rural populations comprised 14% of the participants recruited in November. In December and January, study enrollment shifted to prioritize unvaccinated participants, preferentially pulling these participants from the study waitlist, resulting in 24.6% and 21.4% of monthly enrollment being unvaccinated (Table 1). In January, recruitment efforts also focused on increasing racial and ethnic diversity in the study population, and the minority of participants enrolled during January identified as White (42.6%). Additionally, 10.5% of participants enrolled in January were of Hispanic origin.

**Figure 2:**
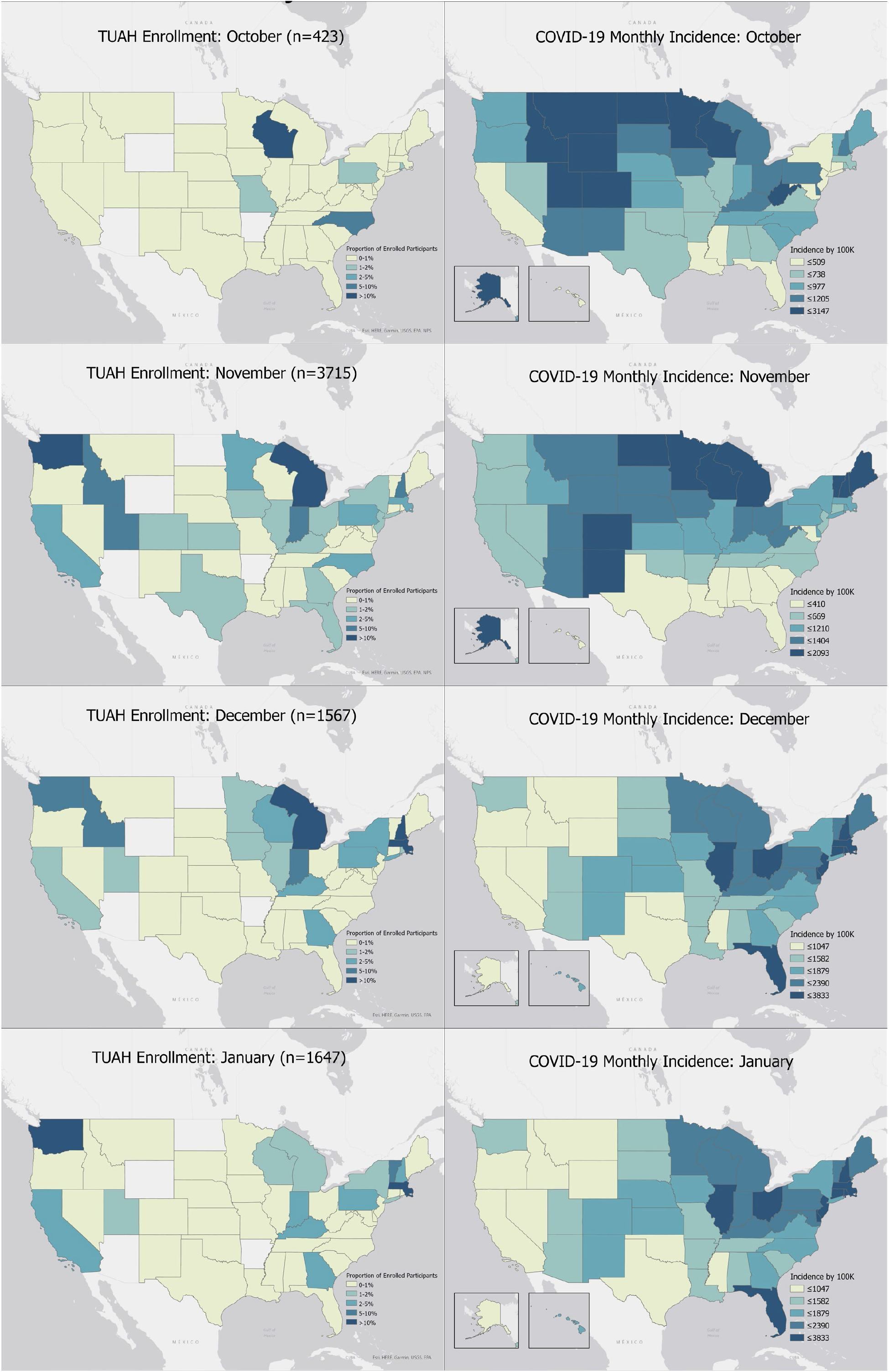
Test Us at Home Enrollment by State and Month, October 2021-January 2022. Note: COVID-19 monthly incidence calculated from CDC COVID Data Tracker^6^

**Figure 3:**
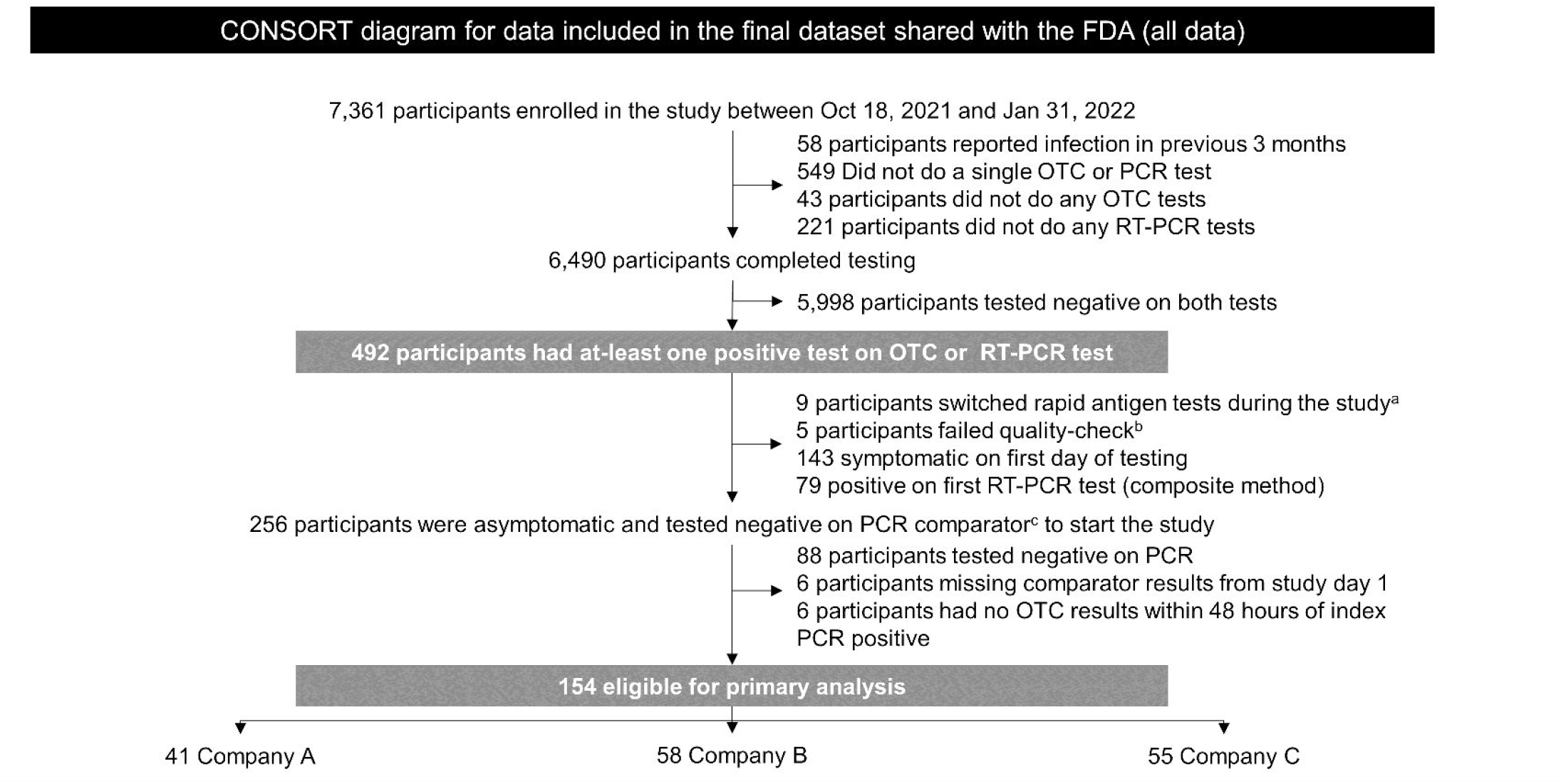
CONSORT diagram of the Test Us At Home Study. a= participants replaced their assigned rapid antigen tests with commercially obtained rapid antigen tests; b= dates of RT-PCR testing could not be verified based on triangulation of self-reported, shipping, and resulting data; A, B, and C refer to rapid antigen test assignment, c = at-least two positive molecular assays from a single sample per participant (Roche Cobas 6800, Quest LDT. Hologic Aptima)

**Table 1:**
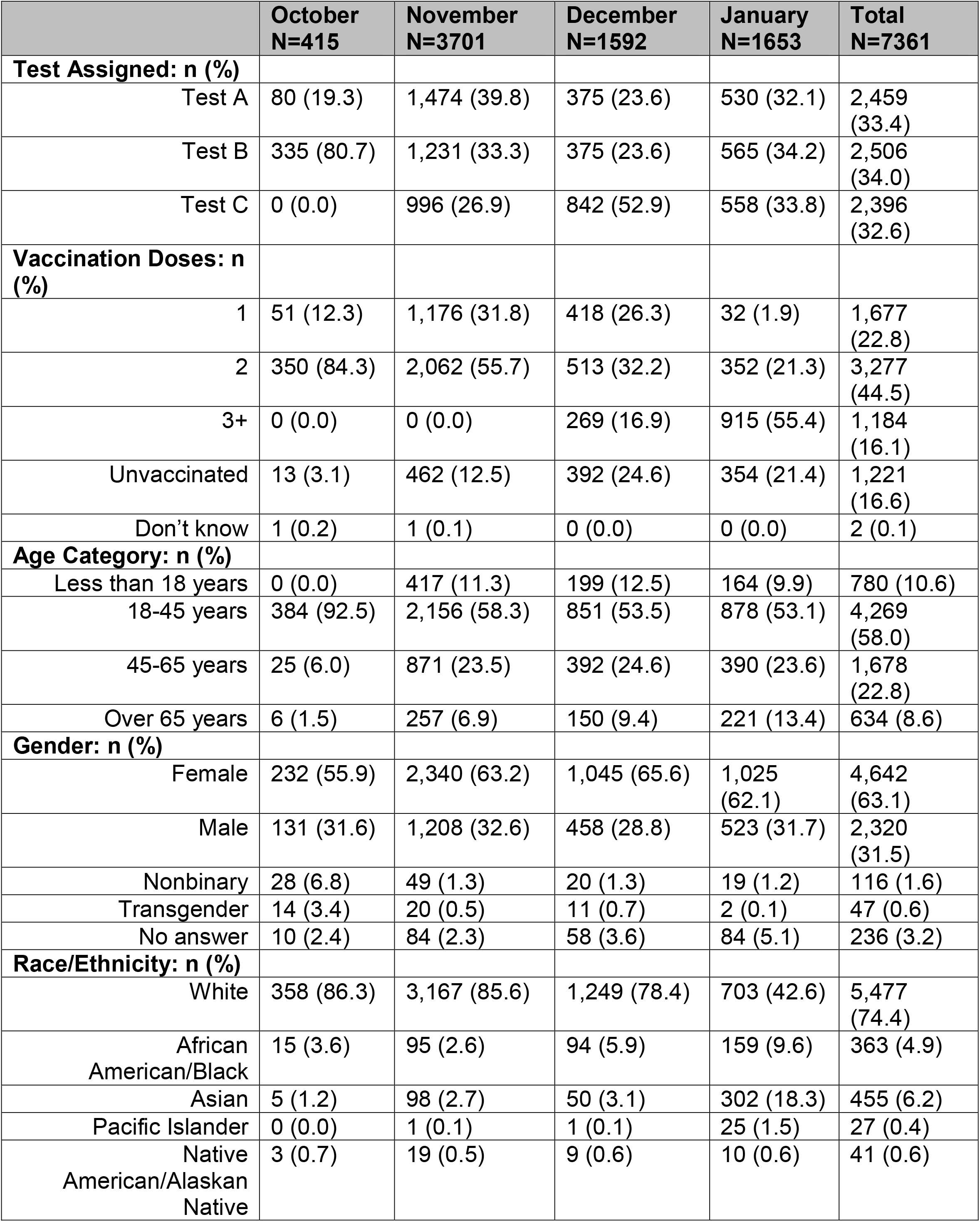
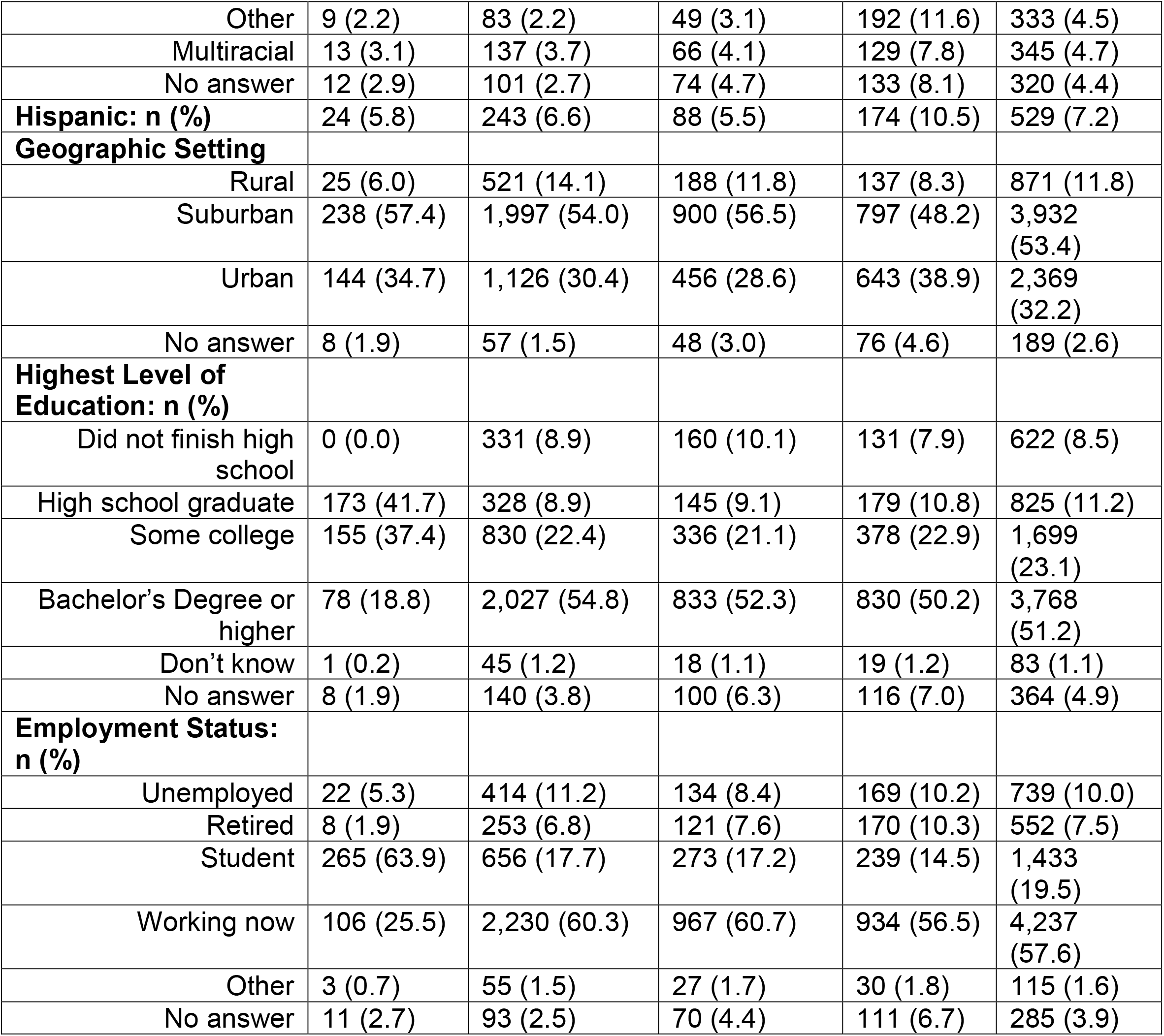
Test Us at Home Participant Demographics by Month of Enrollment.

|  | October<br>N=415 | November<br>N=3701 | December<br>N=1592 | January<br>N=1653 | Total<br>N=7361 |
| --- | --- | --- | --- | --- | --- |
| **Test Assigned: n (%)** |  |  |  |  |  |
| Test A | 80 (19.3) | 1,474 (39.8) | 375 (23.6) | 530 (32.1) | 2,459<br>(33.4) |
| Test B | 335 (80.7) | 1,231 (33.3) | 375 (23.6) | 565 (34.2) | 2,506<br>(34.0) |
| Test C | 0 (0.0) | 996 (26.9) | 842 (52.9) | 558 (33.8) | 2,396<br>(32.6) |
| **Vaccination Doses: n (%)** |  |  |  |  |  |
| 1 | 51 (12.3) | 1,176 (31.8) | 418 (26.3) | 32 (1.9) | 1,677<br>(22.8) |
| 2 | 350 (84.3) | 2,062 (55.7) | 513 (32.2) | 352 (21.3) | 3,277<br>(44.5) |
| 3+ | 0 (0.0) | 0 (0.0) | 269 (16.9) | 915 (55.4) | 1,184<br>(16.1) |
| Unvaccinated | 13 (3.1) | 462 (12.5) | 392 (24.6) | 354 (21.4) | 1,221<br>(16.6) |
| Don't know | 1 (0.2) | 1 (0.1) | 0 (0.0) | 0 (0.0) | 2 (0.1) |
| **Age Category: n (%)** |  |  |  |  |  |
| Less than 18 years | 0 (0.0) | 417 (11.3) | 199 (12.5) | 164 (9.9) | 780 (10.6) |
| 18-45 years | 384 (92.5) | 2,156 (58.3) | 851 (53.5) | 878 (53.1) | 4,269<br>(58.0) |
| 45-65 years | 25 (6.0) | 871 (23.5) | 392 (24.6) | 390 (23.6) | 1,678<br>(22.8) |
| Over 65 years | 6 (1.5) | 257 (6.9) | 150 (9.4) | 221 (13.4) | 634 (8.6) |
| **Gender: n (%)** |  |  |  |  |  |
| Female | 232 (55.9) | 2,340 (63.2) | 1,045 (65.6) | 1,025<br>(62.1) | 4,642<br>(63.1) |
| Male | 131 (31.6) | 1,208 (32.6) | 458 (28.8) | 523 (31.7) | 2,320<br>(31.5) |
| Nonbinary | 28 (6.8) | 49 (1.3) | 20 (1.3) | 19 (1.2) | 116 (1.6) |
| Transgender | 14 (3.4) | 20 (0.5) | 11 (0.7) | 2 (0.1) | 47 (0.6) |
| No answer | 10 (2.4) | 84 (2.3) | 58 (3.6) | 84 (5.1) | 236 (3.2) |
| **Race/Ethnicity: n (%)** |  |  |  |  |  |
| White | 358 (86.3) | 3,167 (85.6) | 1,249 (78.4) | 703 (42.6) | 5,477<br>(74.4) |
| African<br>American/Black | 15 (3.6) | 95 (2.6) | 94 (5.9) | 159 (9.6) | 363 (4.9) |
| Asian | 5 (1.2) | 98 (2.7) | 50 (3.1) | 302 (18.3) | 455 (6.2) |
| Pacific Islander | 0 (0.0) | 1 (0.1) | 1 (0.1) | 25 (1.5) | 27 (0.4) |
| Native<br>American/Alaskan<br>Native | 3 (0.7) | 19 (0.5) | 9 (0.6) | 10 (0.6) | 41 (0.6) |
| Other | 9 (2.2) | 83 (2.2) | 49 (3.1) | 192 (11.6) | 333 (4.5) |
| Multiracial | 13 (3.1) | 137 (3.7) | 66 (4.1) | 129 (7.8) | 345 (4.7) |
| No answer | 12 (2.9) | 101 (2.7) | 74 (4.7) | 133 (8.1) | 320 (4.4) |
| **Hispanic: n (%)** | 24 (5.8) | 243 (6.6) | 88 (5.5) | 174 (10.5) | 529 (7.2) |
| **Geographic Setting** |  |  |  |  |  |
| Rural | 25 (6.0) | 521 (14.1) | 188 (11.8) | 137 (8.3) | 871 (11.8) |
| Suburban | 238 (57.4) | 1,997 (54.0) | 900 (56.5) | 797 (48.2) | 3,932 (53.4) |
| Urban | 144 (34.7) | 1,126 (30.4) | 456 (28.6) | 643 (38.9) | 2,369 (32.2) |
| No answer | 8 (1.9) | 57 (1.5) | 48 (3.0) | 76 (4.6) | 189 (2.6) |
| **Highest Level of Education: n (%)** |  |  |  |  |  |
| Did not finish high school | 0 (0.0) | 331 (8.9) | 160 (10.1) | 131 (7.9) | 622 (8.5) |
| High school graduate | 173 (41.7) | 328 (8.9) | 145 (9.1) | 179 (10.8) | 825 (11.2) |
| Some college | 155 (37.4) | 830 (22.4) | 336 (21.1) | 378 (22.9) | 1,699 (23.1) |
| Bachelor's Degree or higher | 78 (18.8) | 2,027 (54.8) | 833 (52.3) | 830 (50.2) | 3,768 (51.2) |
| Don't know | 1 (0.2) | 45 (1.2) | 18 (1.1) | 19 (1.2) | 83 (1.1) |
| No answer | 8 (1.9) | 140 (3.8) | 100 (6.3) | 116 (7.0) | 364 (4.9) |
| **Employment Status: n (%)** |  |  |  |  |  |
| Unemployed | 22 (5.3) | 414 (11.2) | 134 (8.4) | 169 (10.2) | 739 (10.0) |
| Retired | 8 (1.9) | 253 (6.8) | 121 (7.6) | 170 (10.3) | 552 (7.5) |
| Student | 265 (63.9) | 656 (17.7) | 273 (17.2) | 239 (14.5) | 1,433 (19.5) |
| Working now | 106 (25.5) | 2,230 (60.3) | 967 (60.7) | 934 (56.5) | 4,237 (57.6) |
| Other | 3 (0.7) | 55 (1.5) | 27 (1.7) | 30 (1.8) | 115 (1.6) |
| No answer | 11 (2.7) | 93 (2.5) | 70 (4.4) | 111 (6.7) | 285 (3.9) |

### Operational Support and Logistics

Throughout the study, there were 11,646 contacts between coordinators and 4,389 distinct participants (Supplemental Table 2). This accounted for 33,356 minutes of coordination time. Coordinators and participants communicated by phone call (39.7%), email (30.6%), voicemail (29.7%), and push notification within the study app (4.2%). The most common reason for contact was to share COVID-19 test results from molecular tests, which accounted for 24.3% of all calls. Other common reasons for contact included testing reminders (19.1%), shipping and receipt of test kits (16.7%), compensation (16.3%), and clarifying the testing schedule (10.4%). The company-generated app for Test C was initially only compatible with certain smartphones; therefore, we delayed enrollment for Test C by two weeks and troubleshooted the app with the company, to ensure participants were able to use the app appropriately (Figure 1a). Further, the decentralized design necessitated an increased reliance on shipping vendors, allowing us to reach participants across the mainland US states. However, this also required a variety of tracking methods to keep track of the 58,888 kits shipped. This study spanned the holiday season, which resulted in additional shipping delays; 1.2% (n =728) of packages were affected by logistical or participant errors that led to an unviable or missing RT-PCR test result.

## DISCUSSION

Using an innovative digital, site-less study approach, we developed recruitment and enrollment strategies to capture new onset COVID-19 infections most effectively throughout the country, resulting in the detection of nearly three times as many new-onset COVID-19 cases as originally specified. The digital site-less approach allowed us to seamlessly and dynamically change enrollment patterns and sample different populations based on the evolving nature of the pandemic, to assess rapid diagnostics with agility, efficiency, and rigor among asymptomatic individuals.

The digital site-less approach offered great agility to alter the recruitment strategy to fit our needs throughout the evolution of the study. Initially, to iron out the study logistics, we opened study recruitment in October 2021 to members of a Midwestern university marching band who were traveling together, where we anticipated a high amount of unmasked, close contact interactions. This group enrollment is reflected in the demographics of participants in October, who were younger and had a higher proportion of white participants than participants recruited in November through January. Later, we were able to geographically alter the sampling to enroll participants from throughout the United States and use a waitlist to selectively sample certain populations by zip-code, determined by community prevalence of COVID-19, and demographics to ensure we were curating a representative sample. For example, after seeing low enrollment of participants over age 65 years in October through December, in January, we selectively pulled those over 65 years of age off the waitlist, increasing the proportion of participants in this group from 1.5% in October to 13.4% in January. Additionally, the recruitment strategy resulted in 16.6% of all enrolled participants being unvaccinated, matching nationwide estimates for those over 5 years of age (CDC data tracker). The waitlist approach also allowed us to ensure that enrollment was not solely clustered among those who have greatest access to recruitment strategies, but rather gave the time for information to disseminate to and within various communities.

The remote design also resulted in gains in study efficiency, in terms of both coordinator time and cost. Instead of requiring coordinators to schedule and supervise serial sample collections for each participant, the virtual, site-less design allowed participants to test in their homes and on their time, increasing accessibility to individuals with various employment and school schedules, as well as living situations. According to the study coordination logs, direct participant-coordinator phone calls averaged to just over 1-hour per day (82.3 minutes over 161 days of the study) and consisted primarily of testing reminders and returning molecular test results to participants. As clinical research coordinators on average support upwards of seven studies concurrently and approximately two-thirds of coordinators report being expected to work more time on studies than allotted, it is important to optimize study coordination time, while maintaining quality and consistency.^14^ Here, we showed the ability for site-less digital trials to run with minimal participant-coordinator interaction, while maintaining high enrollment and adherence to the study protocol.

Site-less cohort studies have become increasingly common, as advances in technology have made virtual recruitment and engagement more feasible.^15^ However, digital products and solutions in clinical research remain underutilized.^16^ Site-less studies can not only facilitate recruitment and participant engagement, but they can also enhance scientific rigor and design. Through coordination with shipping services, home-test distributors, and clinical labs, we organized shipments of study supplies to participants on enrollment, which allowed participants to start testing at home within 24 to 48 hours of enrollment. Further, we coordinated the pick-up and shipment of biological samples to the central laboratory, which has seldom been done within the framework of a digital study. The success of these workflows opens the door for the adaptation of site-less studies to answer many complex research questions, by integrating survey responses, biological samples, and home health monitoring.

There are potential limitations to the study. We did not require participants to be under video observation while performing the Ag-RDT or collecting samples for molecular tests. However, our study facilitated use of these tests ‘as intended’ and ‘as authorized’ by the FDA and therefore may represent better approximation of real-world performance of the tests than traditional clinical studies. Additionally, the smartphone app required participants to take images of the Ag-RDT; therefore, we could verify the Ag-RDT results for all participants. Due to the scale and nature of this study, we used commercial vendors for assembly and distribution of the testing kits, which precluded our ability to provide study-specific written instructions enclosed within the kit; we instead relied on providing all study-specific instructions through the smartphone app. This resulted in confusion for some participants who did not start testing immediately after receiving the test kits or did not perform both Ag-RDT and PCR tests concurrently. However, our study team was able to detect these instances and intervene accordingly. Because of the logistical constraints, we could not facilitate a cold-chain transportation of the collected specimen, which preclude our ability to perform viral culture studies. Sample collection for molecular tests required participants to handwrite the date of sample collection, and laboratory data had to be linked with Ag-RDT data based on Participant ID and testing date, which resulted in mismatches and required substantial manual effort for reconciling differences. Finally, the requirement of smartphone ownership to participate in the study disadvantages people without smartphones. However, smartphone use is ubiquitous and has been accelerated by the SARS-CoV-2 pandemic.^17^ Additionally, we submit that the barrier to participation created by requiring a smartphone is lesser than that created by requiring participants to travel to a clinical study site, multiple times during the study.

These limitations notwithstanding, this study represents the most robust attempt at understanding the performance of Ag-RDT for SARS-CoV-2 detection among asymptomatic participants. We summarize the advantages of our digital siteless approach in Table 2 and outline recommendations for future digital studies. The collaboration between the NIH, FDA, and the RADx Clinical Studies Core allowed for development of a protocol that was innovative and a study that will provide tremendous value to federal agencies, academic researchers and companies for elucidating key questions related to raid antigen tests’ performance. Future collaborative efforts to develop best practice guidelines and infrastructure for performing digital clinical studies are needed to advance scientific community’s ability to perform rigorous clinical research and answer vexing questions.

**Table 2.** Summary of features of this study and opportunity for improvement.

| **Challenges** | **Innovation** | **Opportunity for improvement** |
| --- | --- | --- |
| Widespread outreach to asymptomatic individuals | Democratized participation such that anyone, anywhere, anytime can choose to participate through a smartphone without needing to travel to a clinical study site or recruitment centers | Requirement of smartphone to enroll can prevent some people from participating. Approaches that loan study smartphones or leverage community-health workers as digital studies' liaisons can help overcome this limitation |
| Engagement of diverse communities | A dedicated team met with county health workers and community stakeholders from across the country to promote awareness of the study and participation among people of color, indigenous heritage, and rural locations | A network of community partners and stakeholders for digital clinical studies that inform, plan, and conduct outreach efforts can help further enhance participation of historically underrepresented populations |
| Recruitment from SARS-CoV-2 "hotspot" | Use of CDC daily case reports and projects to identify geographical regions where community outreach efforts were enhanced and which were identified as 'preferred regions for enrollment' | Promoting peer-to-peer recruitment from hotspots through the use of the smartphone apps |
| Refinement of enrollment | Adding a layer of prioritization to allow cohort composition such that participants from "hotspots" and those that improve diverse representation of the enrolled cohort | Automatically adaptive priority criteria that does not require manual input or decision-making but can allow manual override |
| Consenting participants including children > 2 years old | Self-contained description of consent, including brief description of key parts of the study displayed through the smartphone app | Use of participatory videos that explain the study objectives and activities embedded within the smartphone app |
| Assigning rapid antigen tests | Pre-specified criteria that assigned tests to participants automatically | Adaptive algorithm that refines criteria to create balanced cohorts based on multiple variables |
| Distributing testing kits | Asynchronous assembly and shipment of kits from commercial vendors | Synchronous assembly and shipment of kits such that all materials arrive in box instead of different boxes |
| Collecting samples for molecular tests | Direct-to-consumer kits for nasal swabs that were returned using FedEx overnight prepaid envelopes | Courier pickup options that can be invoked from within the study app |
| Performing up to three separate molecular assays in CLIA laboratories | Automated ordering of two tests and manual ordering of additional tests when discordance was observed | Automated ordering of discordant testing with streamlined data collection |
| Serial testing within a specific window (44-52 hours apart) | Automated app notification and text messages every two hours during the testing window | Providing the option of defining notification frequency during the testing window to the participants |
| Verification of rapid antigen testing results | In-app widget to allow participants to take an image of rapid antigen tests that were later verified by the coordinators | Automated readers that verify that an image of rapid antigen test was collected and that the test result matches the result manually entered by the participant |
| Data collection and quality | Data fidelity and timestamp for each element collected within the study app and collation of disparate datafeed from the molecular testing laboratory | Minimizing user entry of free-text or handwritten data |
| Participant reimbursement | Batch processing of prepaid Bank of America cards that are mailed to participants' home | Automated delivery of reimbursement through smartphone app once the requirements are met |

## Data Availability

All data produced in the present study are available upon reasonable request to the authors.

## Data Availability

The datasets generated during the current study are available through the RADx Datahub (https://www.radxdatahub.info/).

## Acknowledgment

This study was funded by the NIH RADx Tech program under 3U54HL143541-02S2 and NIH CTSA grant UL1TR001453. The views expressed in this manuscript are those of the authors and do not necessarily represent the views of the National Institute of Biomedical Imaging and Bioengineering; the National Heart, Lung, and Blood Institute; the National Institutes of Health, or the U.S. Department of Health and Human Services. Salary support from the National Institutes of Health U54HL143541, R01HL141434, R01HL137794, R61HL158541, R01HL137734, U01HL146382 (AS, DDM), U54EB007958-13 (YCM, MLR), AI272201400007C, UM1AI068613 (YCM). We are grateful to our study participants and to our collaborators from the National Institute of Health (NIBIB and NHLBI) who provided scientific input into the design of this study and interpretation of our results, but could not formally join as co-authors due to institutional policies and to the Food and Drug Administration (Office of In Vitro Diagnostics and Radiological Health) for their involvement in the primary TUAH study. We received meaningful contributions from Drs. Bruce Tromberg, Jill Heemskerk, Felicia Qashu, Dennis Buxton, Erin Iturriaga, Jue Chen, Andrew Weitz, and Krishna Juluru. We are thankful to the developers of the rapid antigen tests, who donated their tests for research use in this study. We are greatly appreciative of the contribution to this study by the numerous staff at UMass Chan, including critical support from Karen Gilliam, Mary Janet McCarthy, Amber Showers, Cynthia Kinahan, Kimberly Cantin, and Danielle Howard. We would also like to acknowledge the support provided by clinical coordinators from Threewire, Inc. We are thankful to county health departments across the country who helped with recruitment for this siteless study by spreading the word in their networks.

## Competing Interest Statement

VK is principal, and TS, SS, CN, ES, and EH are employees of the health care technology company CareEvolution, which was contracted to configure the smartphone study app, provide operational and logistical support, and collaborate on overall research approach. LS and LR are employees of Quest Diagnostics LLC, which was contracted to provide direct-to-consumer kits, logistical support for nationwide RT-PCR testing, and operational support for producing molecular testing results. DDM reports consulting and research grants from Bristol-Myers Squibb and Pfizer, consulting and research support from Fitbit, consulting, and research support from Flexcon, research grant from Boehringer Ingelheim, consulting from Avania, non-financial research support from Apple Computer, consulting/other support from Heart Rhythm Society. YCM has received tests from Quanterix, Becton-Dickinson, Ceres, and Hologic for research-related purposes, consults for Abbott on subjects unrelated to SARS-CoV-2, and receives funding support to Johns Hopkins University from miDiagnostics. AS receives non-financial support from CareEvolution for collaborative research activities. Additional authors declare no financial or non-financial competing interests.

## Notes

### Author Declarations

The IRB of WIRB-Copernicus Group gave ethical approval for this work (20214875).

### Summary of Updates

Amended authorship to include FDA authors

