## Supplemental Figures for "Finding a Needle in a Haystack: Design and Implementation of a Digital Site-less Clinical Study of Serial Rapid Antigen Testing to Identify Asymptomatic SARS-CoV-2 Infection"

### Supplemental Figure 1: Enrollment process

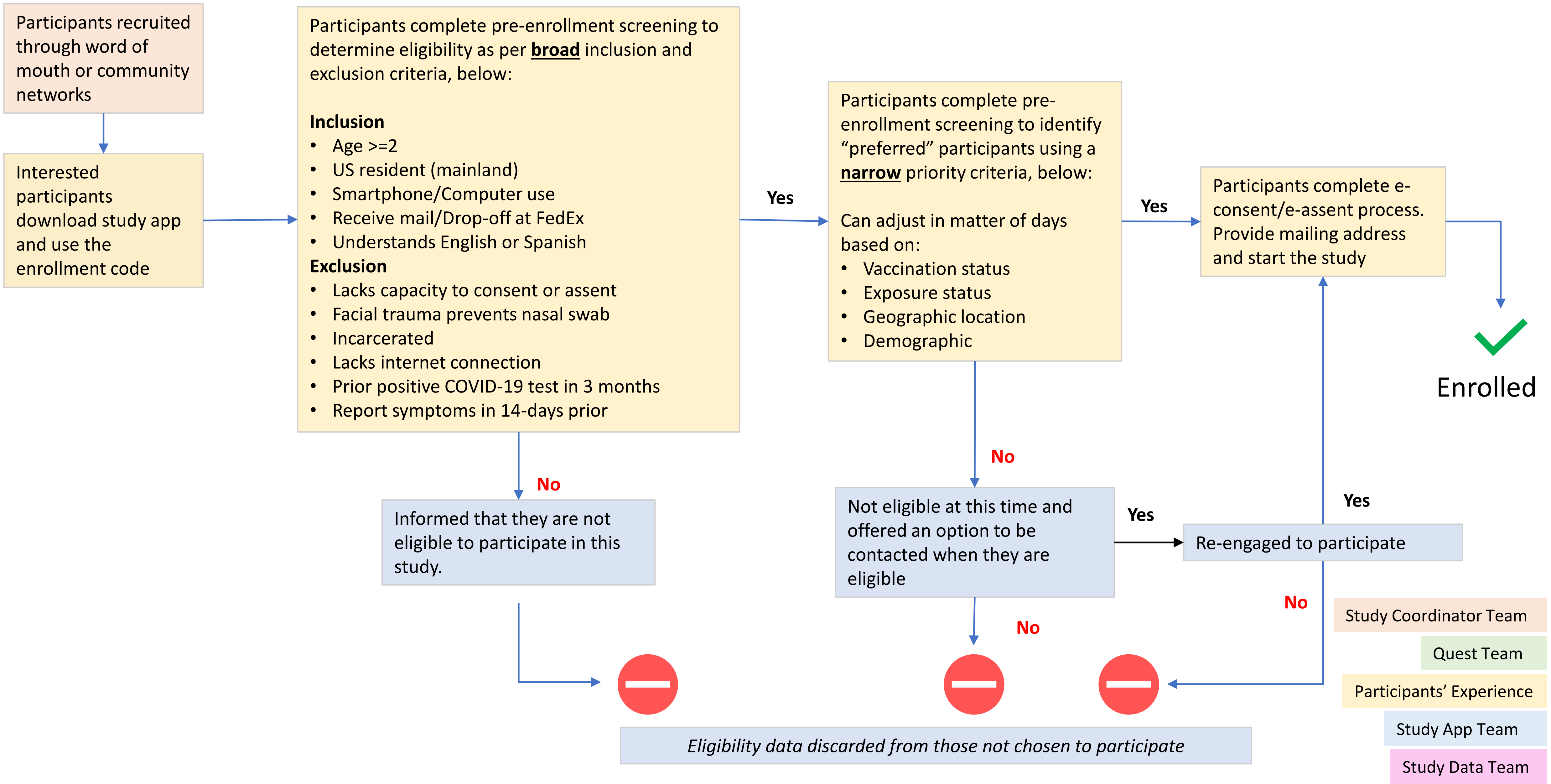

### Supplemental Figure 2: Screenshot of Study App For Determining Preferred Status

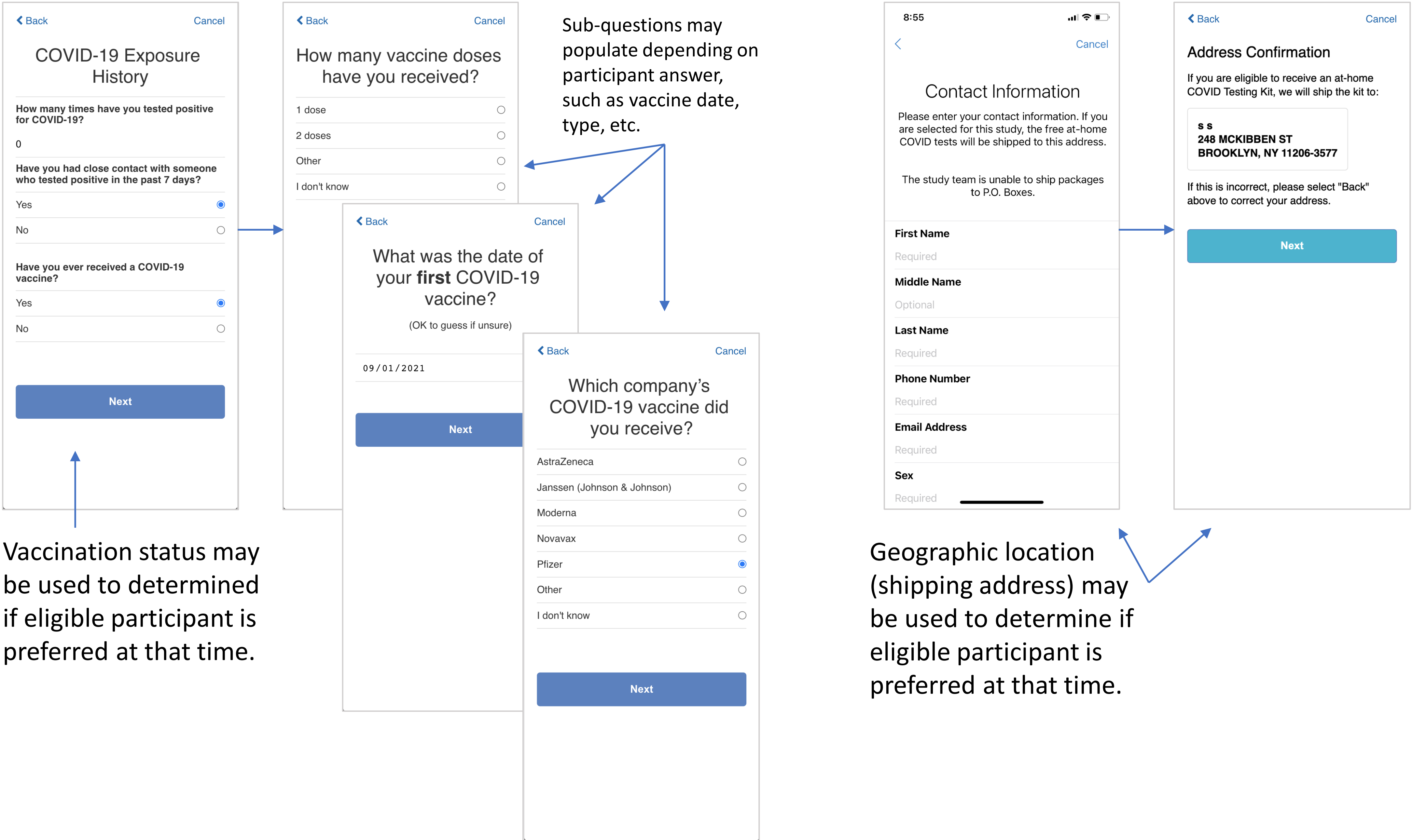


Sub-questions may populate depending on participant answer, such as vaccine date, type, etc.

Vaccination status may be used to determined if eligible participant is preferred at that time.

Geographic location (shipping address) may be used to determine if eligible participant is preferred at that time.

Depending on preferred status criteria at that time, participants will be enrolled or placed on waitlist (Supplemental Figure 3).

### Supplemental Figure 3: Screenshot of Study App For Waitlisted and Preferred Participants

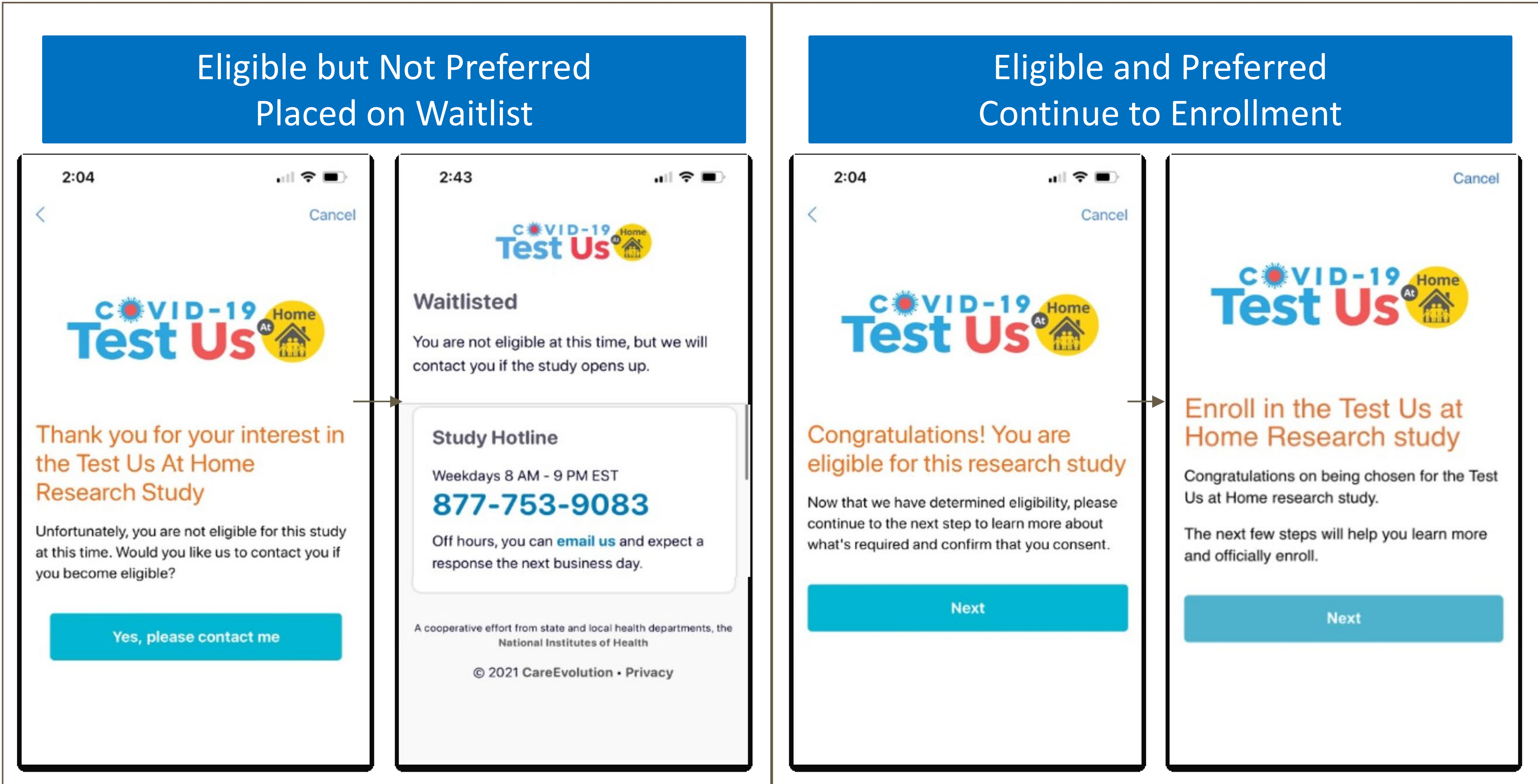

### Supplemental Figure 4: Screenshots of Study App Consent Flow

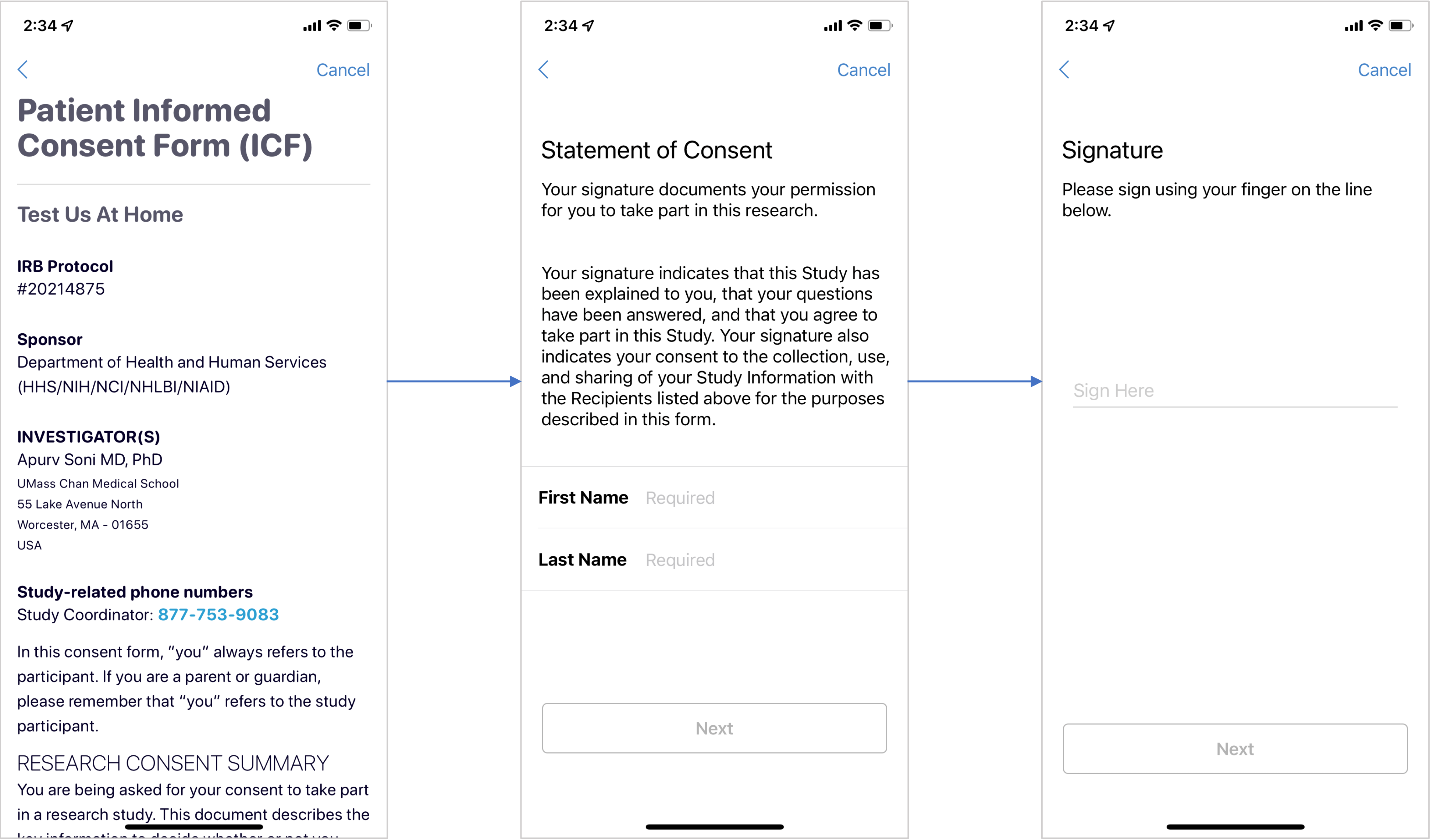

#### Supplemental Figure 5: Participant Compensation

| Criteria | Amount |
|---|---|
| Participants complete 1 OTC test and 1 at home sample collection* on designated test day | $20 per test day (days 1- 13) |
| Participants complete at least 5 OTC tests and at home sample collections* and complete all of the study related activities^ | $30 bonus |
| Participants complete all 8 OTC test and all 7 at home sample collections* and complete all of the study related activities^ | $80 bonus |
| * To be eligible for reimbursement, participant must complete collection and shipping of the sample used for comparator testing (the Quest At Home Collection kit) on the same day of testing with the OTC test if the test is performed on Monday-Thursday; for tests performed on Friday-Sunday, participant must ship the sample on Monday to be eligible for reimbursement.<br><br>^ Study related activities include questionnaires, recording the OTC results, and for some OTC devices uploading an image of the test strip to the study app | |

### Supplemental Figure 6: Test Assignment Strategy

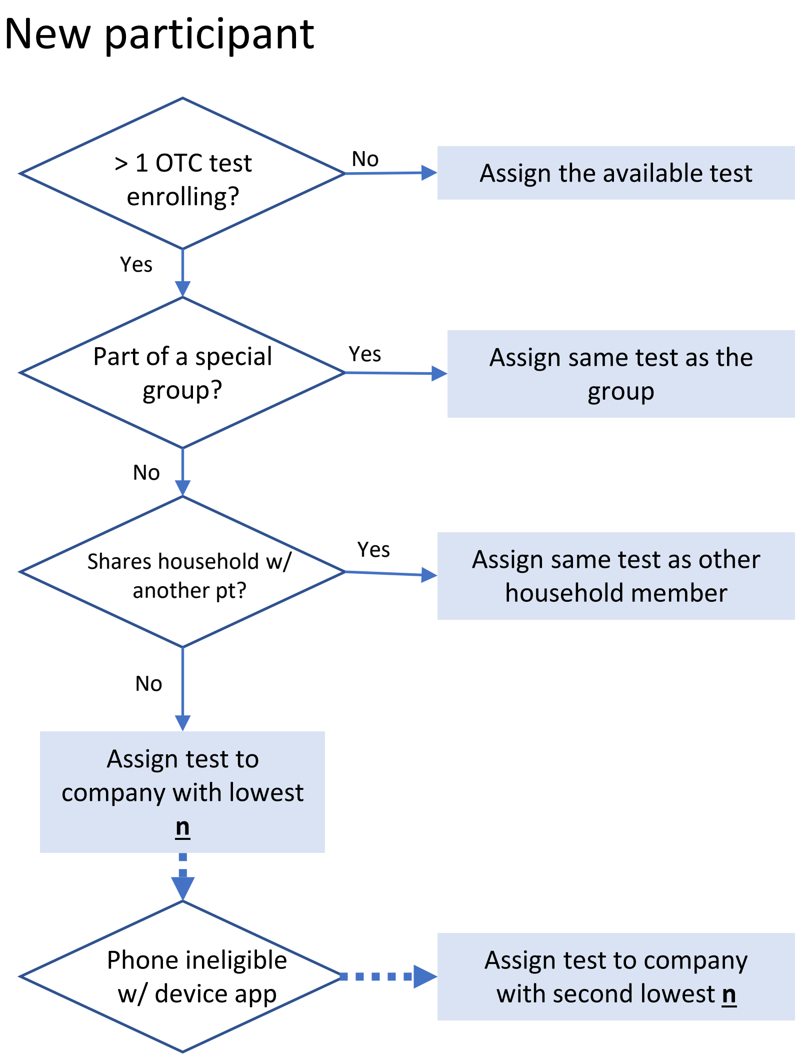


#### Rationale:

For the first week of enrollment, only one device was available. Up to three OTC tests available for OTC test assignment throughout the study.

In order to avoid a case-mix of tests in a specific group, assign participants with the same group code to the same OTC test assignment.

In order to avoid a case-mix of tests in a single household, assign participants with the same street address to the same OTC test assignment.

Algorithm allows the **n** to be based of the following parameters at different timepoints as needed:

- Enrolled
- Asymptomatic positives
- Participants that below 18
- Other characteristics

Study Coordinator Team

Quest Team

Participants' Experience

Study App Team

Study Data Team

### Supplemental Figure 7: Quest Home Kit Orders and OTC Test Delivery

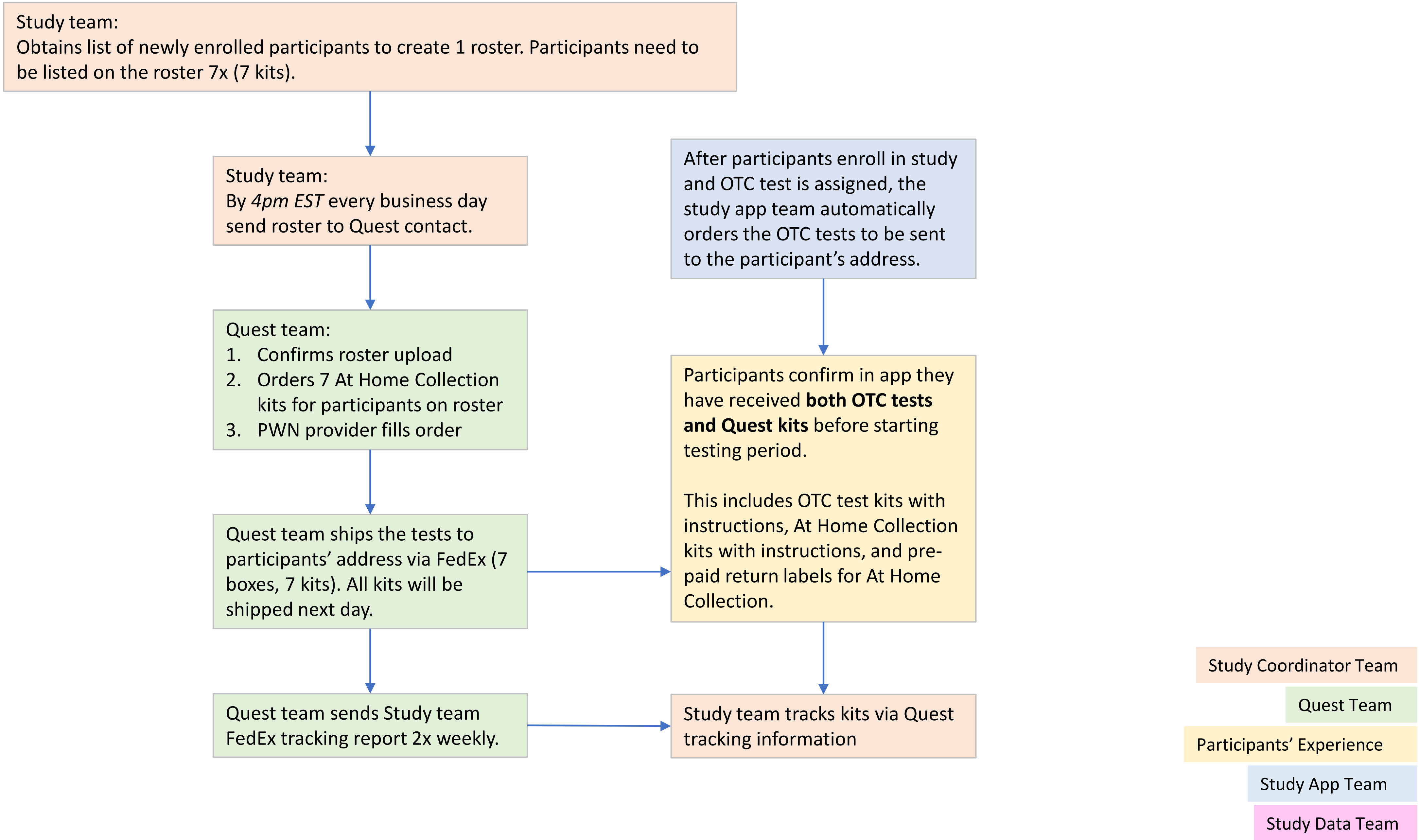

### Supplemental Figure 8: Testing Period and Results

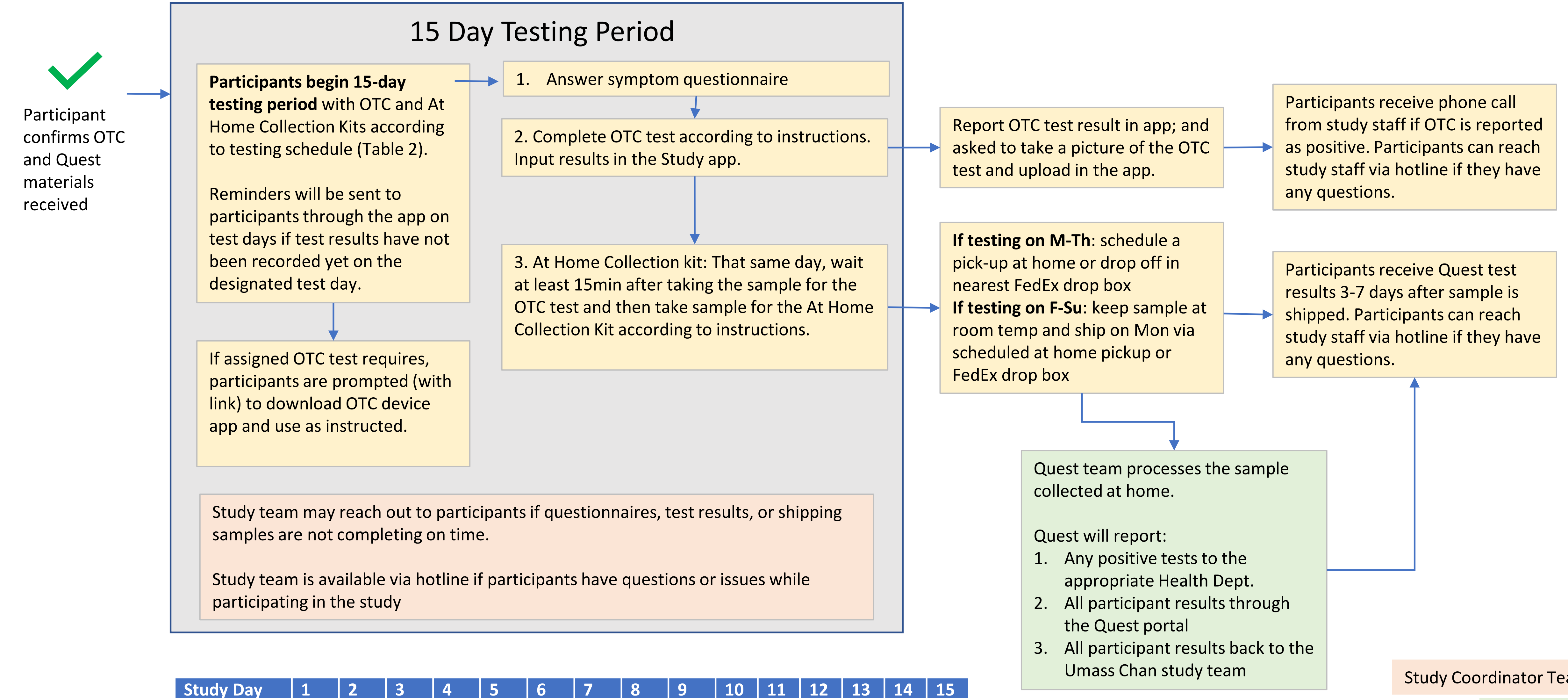


| Study Day | 1 | 2 | 3 | 4 | 5 | 6 | 7 | 8 | 9 | 10 | 11 | 12 | 13 | 14 | 15 |
|---|---|---|---|---|---|---|---|---|---|---|---|---|---|---|---|
| Ag-RDT Device | X | | X | | X | | X | | X | | X | | X | | X |
| Molecular Comparator | X | | X | | X | | X | | X | | X | | X | | |

X=Test completed on this day

Study Coordinator Team
Quest Team
Participants' Experience
Study App Team
Study Data Team

### Supplemental Figure 9: Testing Process

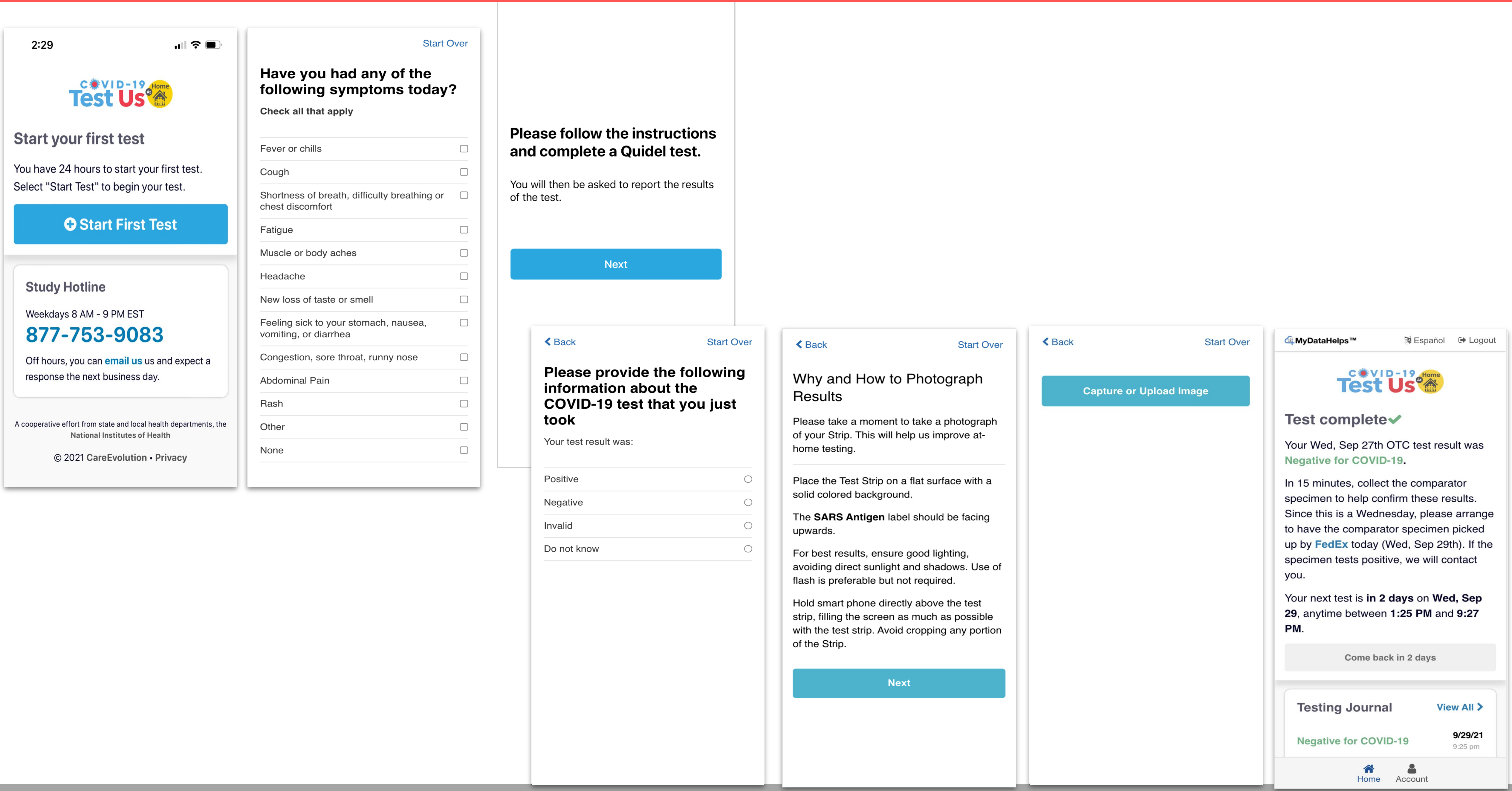

### Supplemental Figure 10: Molecular Comparator Testing and Results

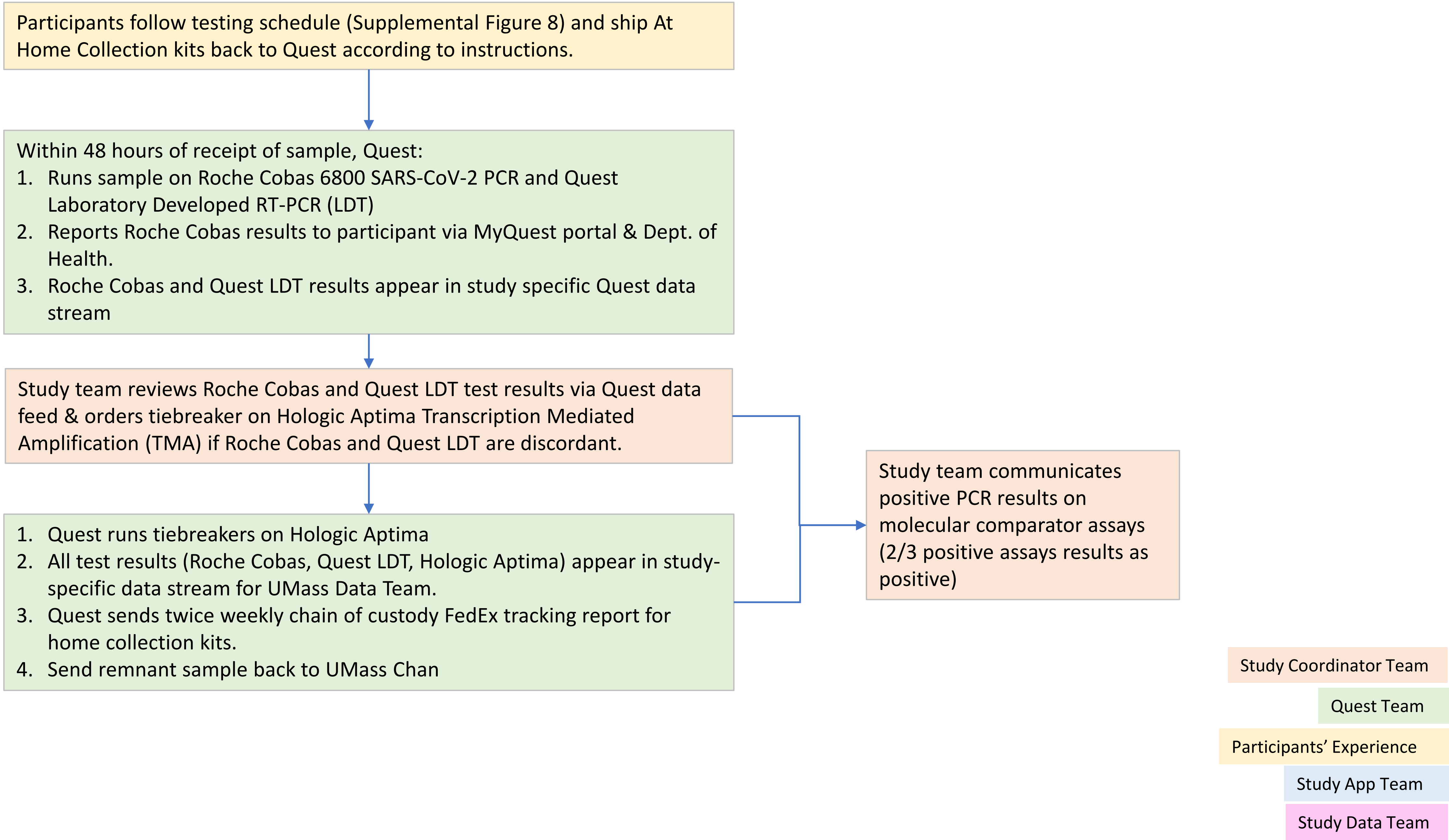

### Supplemental Figure 11: Overview of Test Us At Home Data Flow

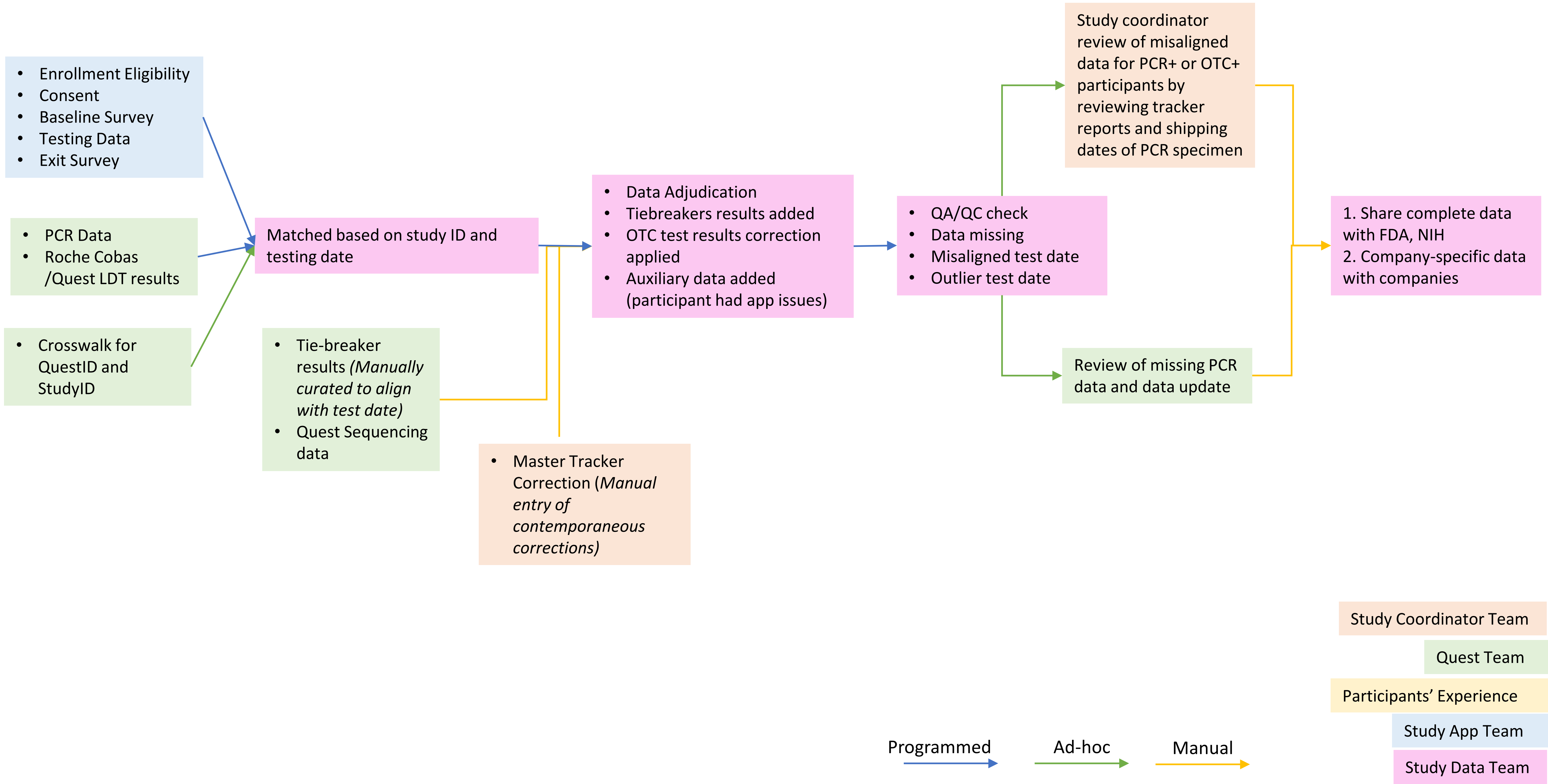

#### Supplemental Figure 12: CONSORT diagram of the Test Us At Home Study

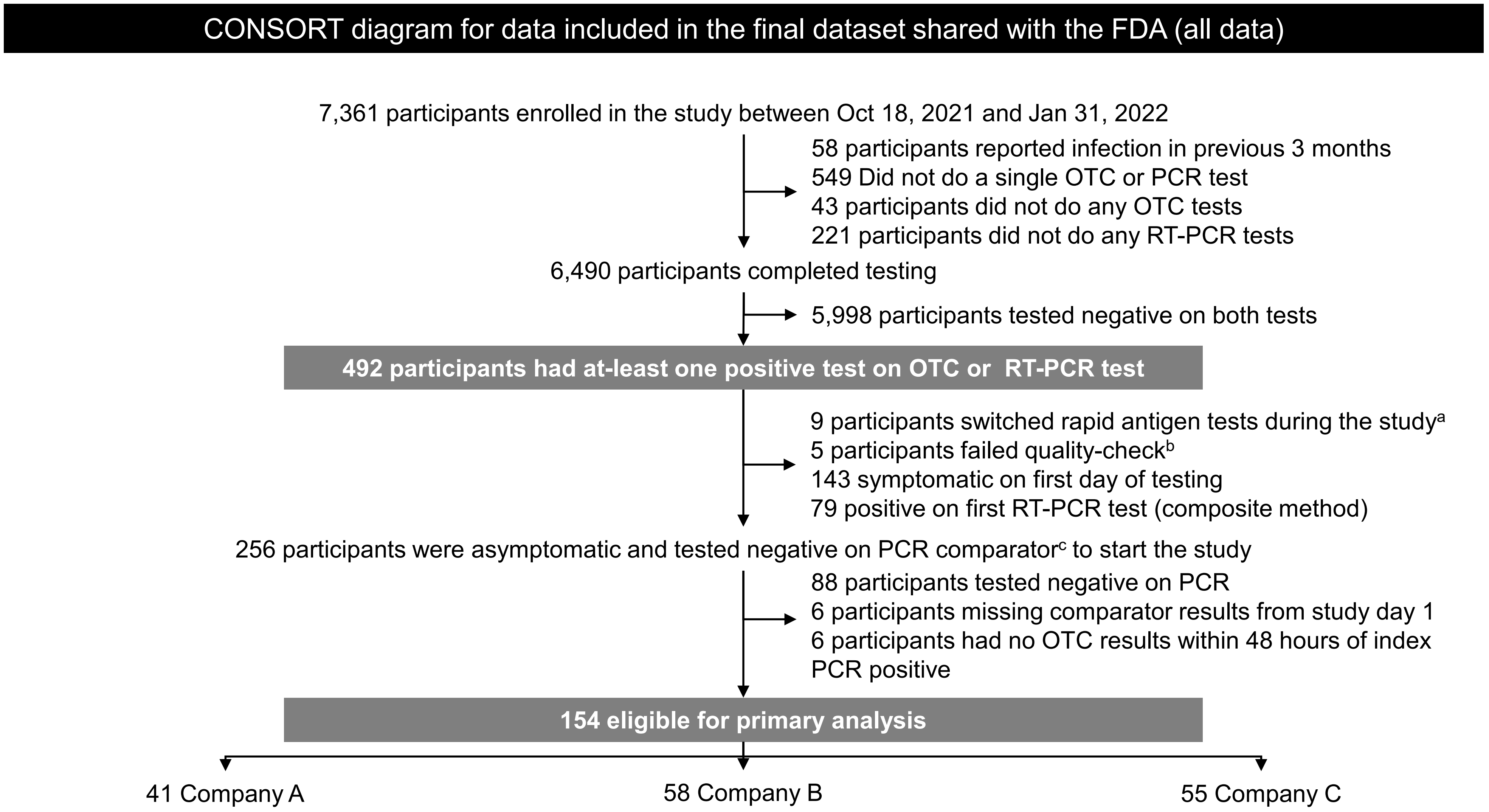


a= participants replaced their assigned rapid antigen tests with commercially obtained rapid antigen tests; b= dates of RT-PCR testing could not be verified based on triangulation of self-reported, shipping, and resulting data; A, B, and C refer to rapid antigen test assignment. c = at-least two positive molecular assays from a single sample per participant (Roche Cobas 6800, Quest LDT, Hologic Aptima)
