## Supplemental Tables for "Finding a Needle in a Haystack: Design and Implementation of a Digital Site-less Clinical Study of Serial Rapid Antigen Testing to Identify Asymptomatic SARS-CoV-2 Infection"

**Supplemental Table 1: Inclusion and Exclusion Criteria**

| Inclusion Criteria |
|---|
| ➢ At least 2 years old<br>➢ Access or parental access to a smartphone or device to download and use study app<br>➢ Reside in mainland United States<br>➢ Able to receive mail at home<br>➢ Willing to drop off comparator samples at FedEx drop-off site or allow for pick-up service to retrieve samples<br>➢ Report no COVID-19 symptoms within 14 days prior to study enrollment |
| Exclusion Criteria |
| ➢ Self-reported positive test for SARS-CoV-2 infection in the previous 3 months<br>➢ Facial trauma interfering with nasal swabs<br>➢ Lack capacity to consent<br>➢ Do not understand English or Spanish<br>➢ Currently in the correctional justice system<br>➢ No internet access on smartphone while at home |

**Supplemental Table 2: Type of Contact and Reasons for Participant-Coordinator Contact**

| | **Number of Participant-Coordinator Contacts** |
|---|---|
| **Total** | **11,646** |
| **Type of Contact, n (%)** | |
| Phone Call | 4,627 (39.7) |
| Voice mail | 3,455 (29.7) |
| Email | 3,564 (30.6) |
| Unscheduled Push notification | 483 (4.2) |
| **Who Initiated Contact?, n (%)** | |
| Participant | 5,371 (46.1) |
| Coordinator | 5,261 (45.2) |
| Both | 825 (7.1) |
| Missing | 189 (1.6) |
| **Reason, n[a]** | |
| App issues | 1,123 (9.6) |
| Interpretation of Test Results | 16 (0.1) |
| Symptom reporting | 63 (0.5) |
| Issues with Rapid Antigen Tests | 251 (2.2) |
| Testing Reminder | 2,228 (19.1) |
| Compensation | 1,903 (16.3) |
| Shipping of Tests | 1,950 (16.7) |
| Study testing schedule | 1,215 (10.4) |
| Giving Test Results | 2,833 (24.3) |
| Other | 1,794 (15.4) |

[a]Reasons for contact were not mutually exclusive
